# Efficacy and tolerability of bevacizumab in patients with severe Covid -19

**DOI:** 10.1101/2020.07.26.20159756

**Authors:** Jiaojiao Pang, Feng Xu, Gianmarco Aondio, Yu Li, Alberto Fumagalli, Ming Lu, Giuseppe Valmadre, Jie Wei, Yuan Bian, Margherita Canesi, Giovanni Damiani, Yuan Zhang, Dexin Yu, Jun Chen, Xiang Ji, Wenhai Sui, Bailu Wang, Shuo Wu, Attila Kovacs, Miriam Revera, Hao Wang, Ying Zhang, Yuguo Chen, Yihai Cao

**Affiliations:** Department of Emergency Medicine, Shandong Provincial Clinical Research Center for Emergency and Critical Care Medicine, Institute of Emergency and Critical Care Medicine of Shandong University, Qilu Hospital of Shandong University, Jinan, Shandong, 250012, China; Clinical Research Center of Shandong University, Jinan, Shandong, 250012, China; Key Laboratory of Cardiovascular Remodeling and Function Research, Chinese Ministry of Education, Chinese National Health Commission and Chinese Academy of Medical Sciences, the State and Shandong Province Joint Key Laboratory of Translational Cardiovascular Medicine, Jinan, Shandong, 250012, China; Department of Medicine and Oncology, Moriggia-Pelascini Hospital, Gravedona ed Uniti, Gravedona, 22015, Italy; Department of Pulmonary and Critical Care Medicine, Qilu Hospital of Shandong University, Jinan, Shandong, 250012, China; Department of Emergency Medicine, Renmin Hospital of Wuhan University, Wuhan, Hubei, 430060, China; Department of Neurological Rehabilitation, Moriggia-Pelascini Hospital, Gravedona ed Uniti, Gravedona, 22015, Italy; Department of Radiology, Moriggia-Pelascini Hospital, Gravedona ed Uniti, Gravedona, 22015, Italy; Department of Radiology, Qilu Hospital of Shandong University, Jinan, Shandong, 250012, China; Department of Radiology, Renmin Hospital of Wuhan University, Wuhan, Hubei, 430060, China; Department of Intensive Care Unit, Moriggia-Pelascini Hospital, Gravedona ed Uniti, Gravedona, 22015, Italy; Department of Cardiology, Moriggia-Pelascini Hospital, Gravedona ed Uniti, Gravedona, 22015, Italy; Department of Critical Care Medicine, Qilu Hospital of Shandong University, Jinan, Shandong, 250012, China; Department of Microbiology, Tumor and Cell Biology, Karolinska Institutet, Stockholm 17177, Sweden

**Keywords:** Covid-19, Bevacizumab, Pneumonia

## Abstract

On the basis of Covid-19-induced pulmonary pathological and vascular changes, we hypothesized that the anti-VEGF drug bevacizumab might be beneficial for treating Covid-19 patients. We recruited 26 patients from 2-centers (China and Italy) with confirmed severe Covid-19, with respiratory rate ≥ 30 times/min, oxygen saturation ≤ 93% with ambient air, or partial arterial oxygen pressure to fraction of inspiration O_2_ ratio (PaO_2_/FiO_2_) >100mmHg and ≤ 300 mmHg, and diffuse pneumonia confirmed by chest radiological imaging. This trial was conducted from Feb 15 to April 5, 2020, and followed up for 28 days. Relative to comparable control patients with severe Covid-19 admitted in the same centers, bevacizumab showed clinical efficacy by improving oxygenation and shortening oxygen-support duration. Among 26 hospitalized patients with severe Covid-19 (median age, 62 years, 20 [77%] males), bevacizumab plus standard care markedly improved the PaO_2_/FiO_2_ ratios at days 1 and 7 (elevated values, day 1, 50.5 [4.0,119.0], *p*<0.001; day 7, 111.0 [85.0,165.0], *p*<0.001). By day 28, 24 (92%) patients showed improvement in oxygen-support status, 17 (65%) patients were discharged, and none showed worsen oxygen-support status nor died. Significant reduction of lesion areas and ratios were shown in chest CT or X-ray analysis within 7 days. Of 14 patients with fever, body temperature normalized within 72 hours in 13 (93%) patients. Lymphocyte counts in peripheral blood were significantly increased and CRP levels were markedly decreased as shown in available data. Our findings suggested bevacizumab plus standard care was highly beneficial for treating patients with severe Covid-19. Clinical efficacy of bevacizumab warrants double blind, randomized, placebo-controlled trials.

**TRIAL REGISTRATION:** ClinicalTrials.gov Identifier: NCT04275414, URL: https://clinicaltrials.gov/ct2/show/NCT04275414.

## INTRODUCTION

Coronavirus disease 2019 (Covid-19) is an ongoing worldwide pandemic.^1^As of July 7, 2020, there were 11,468,979 confirmed cases with 535,181 deaths across 216 countries. Among the confirmed cases, 19%-30% were classified as severe.^2,3^ Dyspnoea caused by inflammatory pulmonary effusion or edema presents in almost all patients with severe Covid-19 and instigates pulmonary and systemic hypoxia.^4-6^ Although many commendable efforts have been made,^7-9^ no drug with demonstrated clinical efficacy for severe Covid-19 is available. Approaches of oxygen-support including mechanical ventilation, non-invasive ventilation, high-flow oxygen, and low-flow oxygen become indispensable, which are closely associated with long hospital stay,^2-4,10^ causing a worldwide shortage of ventilators and other respiratory support medical supplies. The serious pandemic situation poses a global challenge for medical supplies and demands an urgent need for developing effective drugs.^11^

We propose a novel therapeutic concept that blocking vascular endothelial growth factor (VEGF) for treating patients with severe Covid-19 patients. Acute respiratory distress syndrome (ARDS) and dyspnea create hypoxia in lung tissues and other organs. Hypoxia induces VEGF expression through activation of the Prolyl hydroxylases (PHD)-hypoxia inducible factor (HIF)-1 pathway, which upregulates VEGF expression through transcription activation.^12^ VEGF is a potent vascular permeability factor that induces vascular leakiness in Covid-19-infected lung tissues, resulting in plasma extravasation and pulmonary edema, which further increases tissue hypoxia.^13,14^ Additionally, VEGF significantly participates in lung inflammation.^15^ Blocking VEGF and the VEGFR-mediated signaling would improve oxygen perfusion and anti-inflammatory response and alleviate clinical symptoms in patients with severe Covid-19. Thus, we employed bevacizumab, a humanized anti-VEGF monoclonal antibody, for treating patients with severe Covid-19.

Supportive clinical evidence for our therapeutic hypothesis includes: 1) patients with severe Covid-19 suffer from severe hypoxia; 2) VEGF levels in patients with severe Covid-19 are markedly elevated;^5^ 3) pulmonary edema frequently presents in Covid-19 patients;^16,17^ 4) autopsy analysis of Covid-19 patients shows excessive extravasates in alveoli of the infected lungs;^18,19^ 5) vascular disorganization and endothelial cell proliferation in the Covid-19-infected lung tissues,^18,19^ suggesting VEGF-induced vascular effects; 6) overreactive inflammatory response;^20^ and 7) experimental animal models demonstrate that anti-VEGF therapy significantly improves pulmonary edema.^21,22^ In confirmatory with our hypothesis, recent studies have also demonstrated vascular dysfunction of the Covid-19-inefected tissues^23,24^.

Bevacizumab has been used in clinical oncotherapy since 2004, with considerable reliability and safety. Taken together, we designed this trial to investigate clinical benefits of bevacizumab plus standard care for treating patients with severe Covid-19.

## METHODS

### Study Participants

Patients aged 18 to 80 years with a confirmed Covid-19 diagnosis were eligible if they had respiratory distress with a respiratory rate (RR) of ≥ 30 times/min, oxygen saturation (SpO_2_) of ≤ 93% while they were breathing ambient air, or a partial arterial oxygen pressure to fraction of inspiration O_2_ ratio (PaO_2_/FiO_2_) of >100mmHg and ≤ 300 mmHg, and diffuse pneumonia confirmed by chest radiological imaging. A confirmed Covid-19 diagnosis was based on epidemiological history (including cluster transmission) and a positive reverse-transcriptase polymerase-chain-reaction (RT-PCR) assay (BioPerfectus Technologies, China; ELITech Group, France; Seegene, Korea) performed by the local center for disease control or a designated diagnostic laboratory.

The following patients were excluded from the trial: 1) patients with severe hepatic dysfunction (Child-Pugh score ≥ C or aspartate aminotransferase level > 5 times the upper reference limit, URL); 2) patients with severe renal dysfunction (estimated glomerular filtration rate ≤ 30 mL/min/1.73 m^2^) or who required continuous renal replacement therapy, haemodialysis, or peritoneal dialysis; 3) patients with uncontrolled hypertension (sitting systolic blood pressure > 160 mmHg or diastolic blood pressure >100 mmHg) or a history of hypertension crisis or hypertensive encephalopathy; 4) patients with poorly controlled heart diseases, such as New York Heart Association class II or higher cardiac insufficiency, unstable angina pectoris, myocardial infarction within 1 year before enrollment, or supraventricular or ventricular arrhythmia needing treatment or intervention; 5) patients with hereditary bleeding tendency or coagulopathy, and patients who received full-dose anticoagulant or thrombolytic therapy within 10 days before enrollment, or non-steroidal anti-inflammatory drugs with platelet suppression within 10 days before enrolment (except those who used small doses of aspirin [≤ 325 mg/day] for preventive use); 6) patients with thrombosis within 6 months before enrollment, patients who had experienced arterial/venous thromboembolic events, such as ischemic stroke, transient ischemic attack, deep venous thrombosis, or pulmonary embolism within 1 year prior to trial enrollment, and patients with severe vascular diseases (including aneurysms or arterial thrombosis requiring surgery) within 6 months before trial enrollment; 7) patients with unhealed wounds, active gastric ulcers, or fractures; patients with gastrointestinal perforation, gastrointestinal fistula, abdominal abscess, or visceral fistula formation within 6 months before trial enrollment; 8) patients who had undergone major surgery (including preoperative chest biopsy) or received major trauma (such as a fracture) within 28 days before enrollment; patients who might need surgery during the trial; 9) patients with severe, active bleeding such as haemoptysis, gastrointestinal bleeding, central nervous system bleeding, and epistaxis within 1 month before trial enrollment; 10) patients with malignant tumours within 5 years before trial enrollment; 11) patients allergic to bevacizumab or its components; 12) patients with untreated active hepatitis or HIV-positive patients; pregnant and lactating women and those planning to get pregnant; 13) patients who participated in other clinical trials or not considered suitable for this trial by the researchers; or 14) patients who did not provide signed informed consent.

### Control Cohort

An external comparison cohort were selected from the same centers, who met the inclusion criteria as well as the exclusion criteria without severe hepatic, renal or cardiac dysfunction, and are comparable to the bevacizumab-treated groups considering race, age, gender, and the baseline PaO_2_/FiO_2_. The informed consent was waived.

### Trial Design and Oversight

This was a single-arm trial (ClinicalTrials.gov NCT04275414) conducted from February 15 to April 5, 2020 (enrollment of the last patient), which enrolled patients in Renmin Hospital of Wuhan University, Wuhan, Hubei Province, China from February 15 to March 8, 2020, and enrolled patients in Italia Hospital S.p.A. Ospedale Generale di Zona Moriggia - Pelascini, Gravedona ed Uniti (CO), Italy from March 25 to April 5, 2020, and followed up for 28 days or until hospital discharge. All eligible patients evaluated by investigators were enrolled. Each eligible patient received a single dose (500 mg) of bevacizumab (Qilu Pharmaceutical Co. LTD and Roche Pharmaceutical Co. LTD) dissolved in 100 ml of saline intravenously in no less than 90 min under electrocardiography monitoring and standard care. Standard care included supplemental oxygen, non-invasive and invasive ventilation, antivirotic or antibiotic agents, vasopressor support, and extracorporeal membrane oxygenation as necessary. The trial was approved by the ethics committees of Qilu Hospital of Shandong University, Renmin Hospital of Wuhan University, and Italia Hospital S.p.A. Ospedale Generale di Zona Moriggia - Pelascini, Gravedona ed Uniti (CO). Written informed consents were obtained from all patients or the legal representatives of patients if they were unable to provide consents by themselves. The adverse events were monitored and adjudicated by the Safety Monitoring Committee. All the adverse events were handled timely with proper medical treatment to avoid further damage. The trial was conducted in accordance with the principles of the Declaration of Helsinki and the Good Clinical Practice guidelines of the International Conference on Harmonisation.

### Outcomes and Measures

Primary outcomes were changes of PaO_2_/FiO_2_ at day 1 and day 7. PaO_2_ was measured by arterial blood gas (ABG) and FiO_2_ at the time of clinical ABG was obtained. Secondary outcomes included change of chest radiological imaging at day 7, as well as oxygen-support status, discharge rate and change of fever symptom during 28 days follow-up. Oxygen-support status referred to mechanical ventilation, non-invasive ventilation, a transition status of alternate use of non-invasive ventilation and high-flow oxygen, high-flow oxygen, low-flow oxygen or ambient air. Chest radiological imaging referred to chest CT or X-ray, which was quantified or semi-quantified by software or team of experts, respectively. Fever was defined as axillary temperature ≥ 37.5°C.

### Disease Severity Classification

The severity of Covid-19 was defined according to the Chinese Clinical Guidance for Covid-19 Diagnosis and Treatment (the 7th edition). According to this guidance, Covid-19 patients were triaged into the following categories: 1) severe type was defined as adults meeting any of the following criteria: a. shortness of breath, RR≥ 30 times/min; b. oxygen saturation SpO2 ≤ 93% at rest; c. PaO_2_/FiO_2_≤ 300mmHg, patients whose pulmonary imaging displays significant progression of lesion>50% within 24-48 hours should be treated as severe type; 2) critically severe type, patients met any of the following: a. respiratory failure requiring mechanical ventilation; b. shock; c. critical comorbidity with other organ failure that needs ICU monitoring and treatment.

### Clinical, Laboratory and Radiological Data Collection

Arterial blood gas analysis, chest computed tomography (CT) scanning, chest X-ray, and laboratory tests were performed in Renmin Hospital of Wuhan University, and Moriggia – Pelascini Hospital respectively. The patients’ PaO_2_/FiO_2_ ratios were assessed at baseline (within 24 hours prior to bevacizumab administration), on days 1 and 7; chest CT were performed at baseline (within 48 hours prior to bevacizumab administration) and day 7 (±1 day), or alternatively performed chest X-ray at baseline (within 48 hours prior to bevacizumab administration) at days 3 and 7; laboratory tests including blood routine and C-reactive protein (CRP) at baseline (within 48 hours prior to bevacizumab administration) and day 7 was added into the protocol conducted in the Italian population after trial commencement, because we observed a phenomenon of rapid abatement of fever post-bevacizumab administration in the middle of the study. Clinical data including demographic data, presenting symptoms, the changes of status of oxygen-support as well as the symptom of fever were recorded using case record forms. This information was sequently entered into an electronic database and validated by trial staff. Chest CT images were quantified using a medical imaging diagnosis support platform (Huiying Medical Technology Co. LTD, Beijing) to obtain the volume and the ratios of the lesions in bilateral lungs. X-ray images were semi-quantified by the Committee of Imaging Experts using the above platform to assess the ratios of the lesions in bilateral lungs.

### Statistical Analysis

No sample-size calculations were performed. The population under analysis included eligible patients who received a single dose of bevacizumab, and for whom clinical data were available. Categorical variables were reported as numbers and percentages, and quantitative variables as means and standard deviations or medians and quartiles. To compare differences between different time points after intervention and baseline point, a paired t-test or Wilcoxon matched-pairs signed-rank test was used for quantitative data. Statuses of oxygen-support and body temperature during 28 days in the bevacizumab-treated population were described with Gantt charts. Safety measurements applied to all patients exposed to bevacizumab. Statistical significance was indicated by a *p* value of 0.05 and was determined with the use of a two-sided hypothesis test. Wilcoxon signed-rank test and Chi-square test were used to compare the external control and bevacizumab group. All analyses were conducted with SAS software, version 9.4 (SAS Institute, USA).

## RESULTS

### Study Participants

Twenty-seven patients were assigned to receive a single dose of bevacizumab, including 13 patients from China and 14 patients from Italy, among whom 1 patient dropped out owing to withdrew consent (Fig. 1). The median age of patients was 62 years, and 20 (77%) were men. The median time from symptom onset to admission were 10 days, while the median interval from admission to bevacizumab administration were 7 days. Fever 25 (96%), dry cough 21 (81%), shortness of breath 19 (73%), and fatigue 17 (65%) were common onset symptoms. Half of the patients had hypertension history and six (23%) had diabetes history. Patients in Chinese and Italian groups were similar in age, maximum body temperature, medical history, and common clinical symptoms. Days from hospitalization to bevacizumab treatment were significantly longer for patients in the Chinese group than the Italian group (Table 1).

**Table 1.**
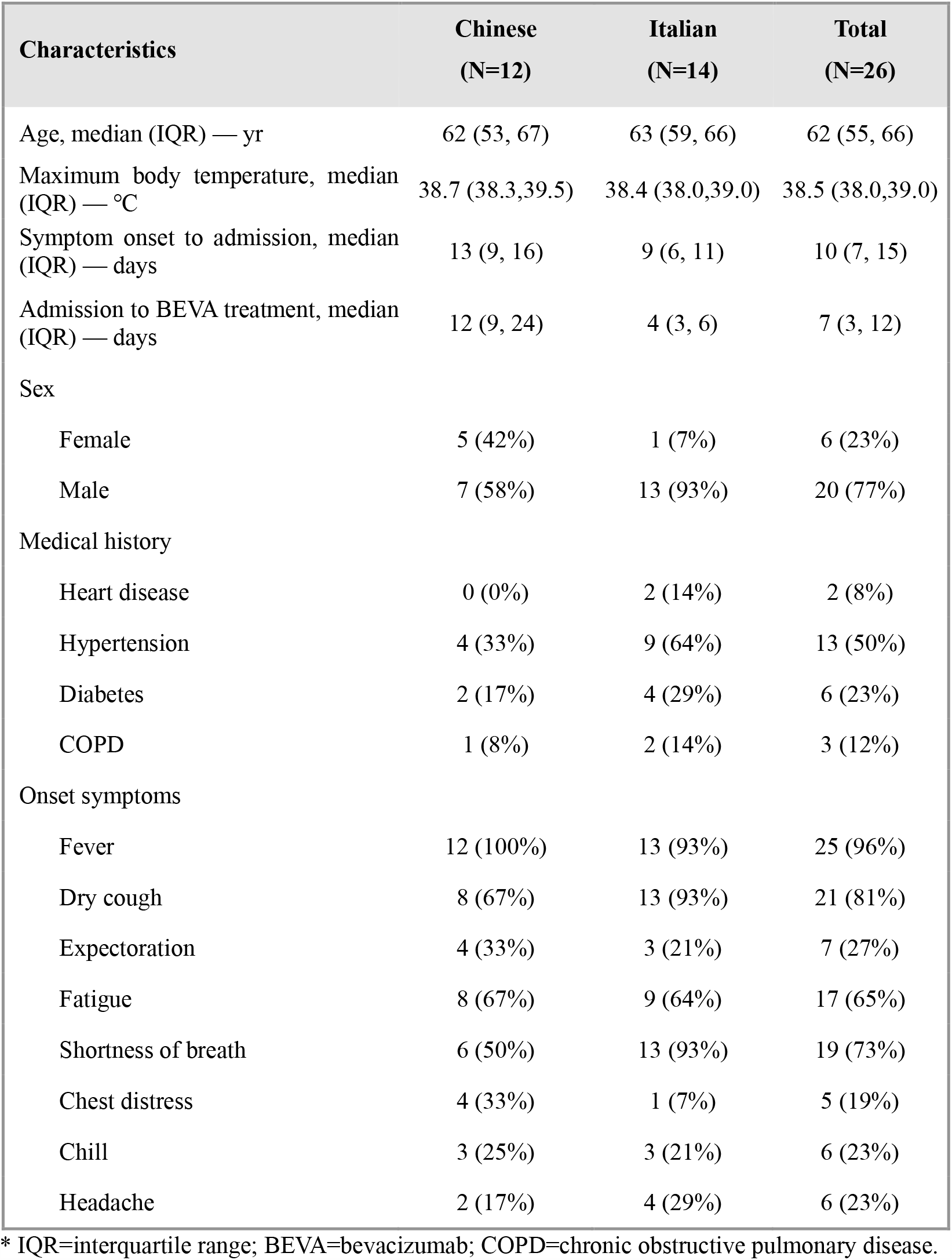
Baseline demographic and clinical characteristics of the patients*

| Characteristics | Chinese<br>(N=12) | Italian<br>(N=14) | Total<br>(N=26) |
| --- | --- | --- | --- |
| Age, median (IQR) — yr | 62 (53, 67) | 63 (59, 66) | 62 (55, 66) |
| Maximum body temperature, median (IQR) — °C | 38.7 (38.3,39.5) | 38.4 (38.0,39.0) | 38.5 (38.0,39.0) |
| Symptom onset to admission, median (IQR) — days | 13 (9, 16) | 9 (6, 11) | 10 (7, 15) |
| Admission to BEVA treatment, median (IQR) — days | 12 (9, 24) | 4 (3, 6) | 7 (3, 12) |
| Sex |  |  |  |
| Female | 5 (42%) | 1 (7%) | 6 (23%) |
| Male | 7 (58%) | 13 (93%) | 20 (77%) |
| Medical history |  |  |  |
| Heart disease | 0 (0%) | 2 (14%) | 2 (8%) |
| Hypertension | 4 (33%) | 9 (64%) | 13 (50%) |
| Diabetes | 2 (17%) | 4 (29%) | 6 (23%) |
| COPD | 1 (8%) | 2 (14%) | 3 (12%) |
| Onset symptoms |  |  |  |
| Fever | 12 (100%) | 13 (93%) | 25 (96%) |
| Dry cough | 8 (67%) | 13 (93%) | 21 (81%) |
| Expectoration | 4 (33%) | 3 (21%) | 7 (27%) |
| Fatigue | 8 (67%) | 9 (64%) | 17 (65%) |
| Shortness of breath | 6 (50%) | 13 (93%) | 19 (73%) |
| Chest distress | 4 (33%) | 1 (7%) | 5 (19%) |
| Chill | 3 (25%) | 3 (21%) | 6 (23%) |
| Headache | 2 (17%) | 4 (29%) | 6 (23%) |

\* IQR=interquartile range; BEVA=bevacizumab; COPD=chronic obstructive pulmonary disease.

**Figure 1.**
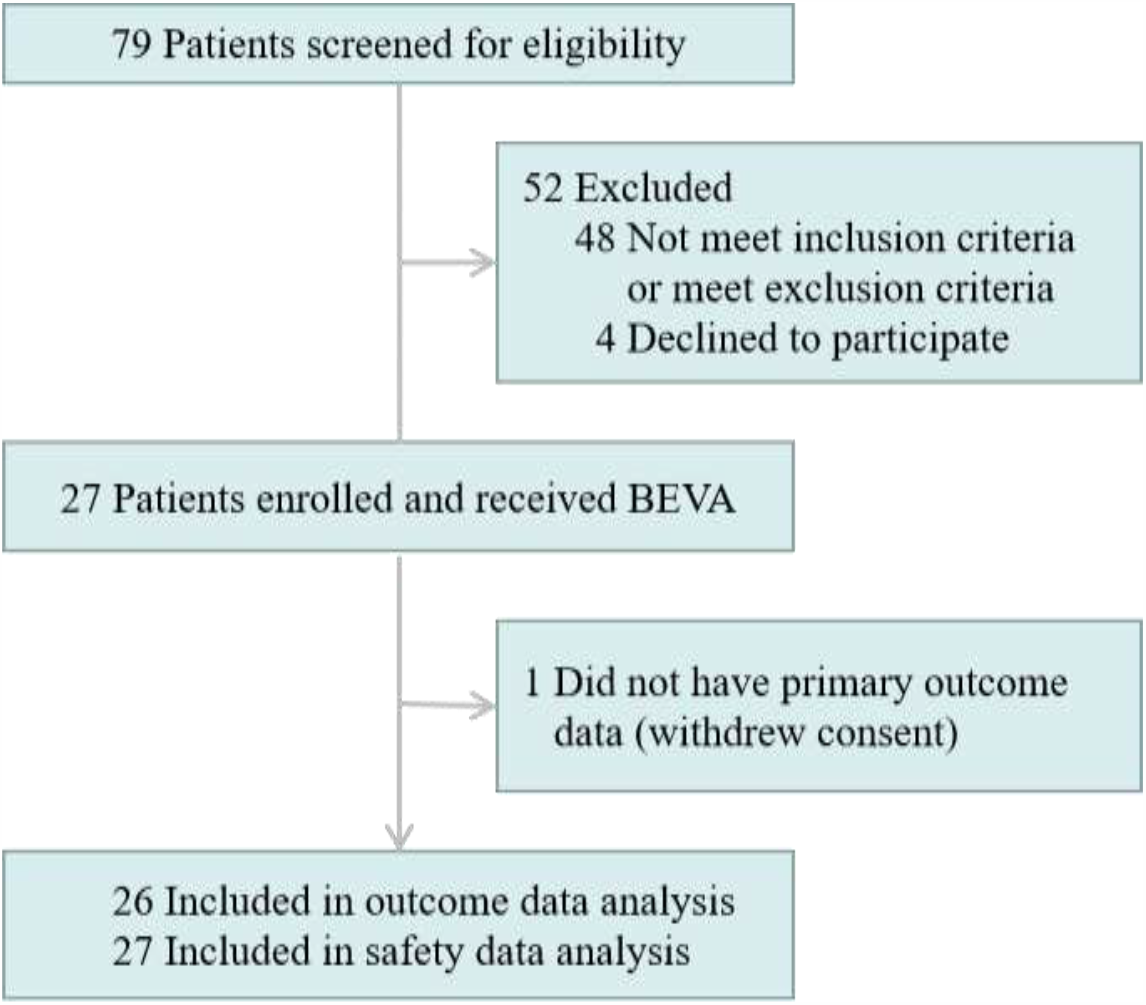
CONSORT flow diagram.

### PaO_2_/FiO_2_ Ratios

PaO_2_/FiO_2_ values markedly increased at days 1 and 7 after bevacizumab administration compared to the baseline values. Differences in genetic background, treatment approaches, and admission-to-bevacizumab administration time between the Chinese and Italian populations should not be ignored. Therefore, we performed a subgroup analysis. Both populations showed markedly increases of PaO_2_/FiO_2_ values at day 1 post-bevacizumab treatment relative to the baseline values, whereas at day 7, only Italian population displayed increases of PaO_2_/FiO_2_ ratios with statistical significance. Among all the patients, 20 of the 26 patients (77%) showed improvement of PaO_2_/FiO_2_ ratios after bevacizumab treatment (Table 2).

**Table 2.** PaO_2_/FiO_2_ ratios*

| Groups | Baseline |  | Difference between day 1 and baseline |  | Difference between day 7 and baseline |  | Number (%) of patients with improved PaO<sub>2</sub>/FiO<sub>2</sub> ratios |
| --- | --- | --- | --- | --- | --- | --- | --- |
|  | N | median (IQR) | median (IQR) | *p*<sup>†</sup> value | median (IQR) | *p*<sup>†</sup> value |  |
| All | 26 | 196.5<br>(145.0,273.0) | 50.5<br>(4.0,119.0) | <0.001 | 111.0<br>(85.0,165.0) | <0.001 | 20 (77%) |
| Chinese group | 12 | 275.0<br>(231.0,339.5) | 68.5<br>(7.5,139.0) | 0.006 | 93.0<br>(-9.0,201.0) | 0.054 | 9 (75%) |
| Italian group | 14 | 151.5<br>(140.0,181.0) | 35.5<br>(4.0,95.0) | 0.004 | 120.5<br>(90.0,165.0) | <0.001 | 11 (79%) |

\*PaO<sub>2</sub>/FiO<sub>2</sub>=partial arterial oxygen pressure to fraction of inspiration O<sub>2</sub> ratio.
IQR=interquartile range. One datum was absent at day 7 because of discharge;
†Wilcoxon matched-pairs signed-rank test.

### Oxygen-Support Status

After receiving a single dose of bevacizumab, 24 of 26 patients (92%) showed marked improvement and 2 patients (8%) showed no change in oxygen-support within 28-day follow-up. Inspiringly, 17 of 26 patients (65%) were discharged. Six patients receiving invasive or non-invasive ventilation stopped ventilator support and 4 of 6 patients (67%) were discharged within follow-up. For the 2 patients with no change in oxygen-support, one breathed ambient air at baseline. None of the 26 patients showed worsened status of oxygen-support nor died after bevacizumab treatment in the 28-day follow-up trial (Fig. 2).

**Figure 2.**
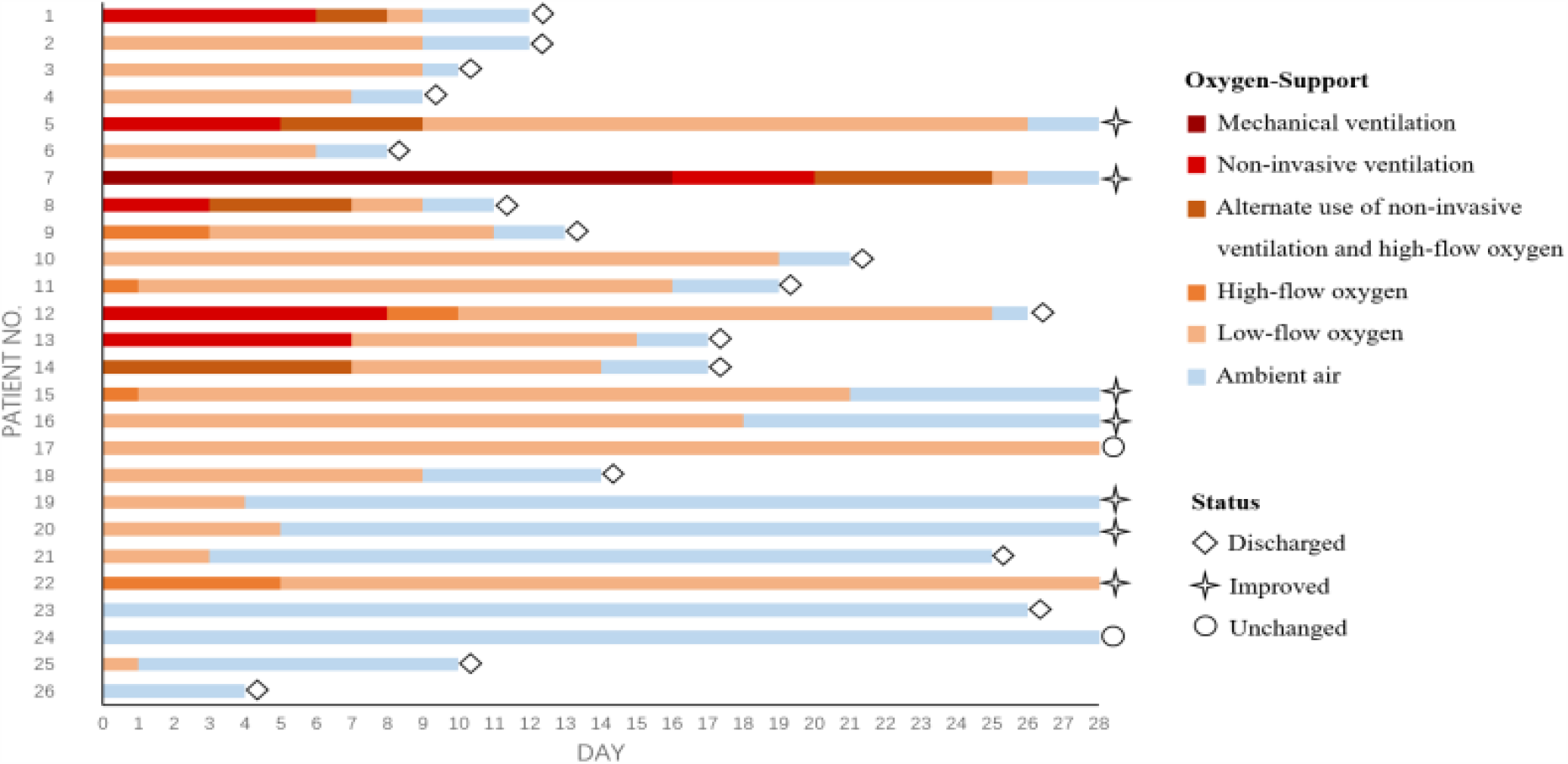
Changes in oxygen-support status in individual patients. Baseline (day 0) is the day when treatment with a single dose of bevacizumab was performed. The final oxygen-support statuses are on the discharge date or at the end of 28-day follow-up. For each individual patient, colored columns represent the oxygen-support status of the patient over time. Diamond symbols represent patients discharged from hospitals; star symbols represent patients improved but not discharged; and circle symbols show patients unchanged.

### Chest Radiological Imaging

Eight patients received chest CT scanning and 12 patients received bedside or traditional X-ray examination alternatively at the required timepoints. Six patients, despite of good recovery (patient No. 15, 19, 21, 22, 23, and 26 shown in Fig. 2), were unable to receive chest radiological examination in time due to unavailability of CT machines, shortage of medical staff, or being discharged. The quantified results of chest CT showed that the total lesion areas (cm^3^) and the lesion ratios (%) for both the lungs significantly reduced at day 7 relative to baseline. Regarding changes in characteristic pulmonary lesions, the number of ground-glass opacities increased and the number of patchy shadows decreased at day 7 compared to the baseline value, which indicated the pulmonary inflammatory exudation was progressively absorbed, however the increase and decrease were not significant. No consolidation lesions were shown in chest CT images of these patients. The semi-quantification of chest X-ray images showed, the lesion ratios of the right lungs remarkably reduced at day 3 and day 7 compared to baseline (Table 3). The representative chest CT as well as X-ray images were shown in Fig.3 and Fig.4.

**Table 3.** Chest radiological imaging quantification*

| Chest CT quantification | N | Baseline | Day 3 |  | Day 7 |  |
| --- | --- | --- | --- | --- | --- | --- |
|  |  | Median (IQR) | Median (IQR) | *p*<sup>†</sup> value | Median (IQR) | *p*<sup>†</sup> value |
| Patchy shadow | 8 | 2.5 (0-4.0) | -- | -- | 1.5 (0-3.5) | 0.641 |
| Ground-glass opacity | 8 | 3.5 (1.5-7.0) | -- | -- | 6.5 (3.5-10.5) | 0.094 |
| Total Lesion area (cm<sup>3</sup>) | 8 | 1055.9 (532.8-1409.4) | -- | -- | 680.7 (395.9-926.2) | 0.039 |
| Lesion ratio (%)<br>— Right Lung | 8 | 19.2 (12.8-33.6) | -- | -- | 12.4 (9.7-15.6) | 0.023 |
| Lesion ratio (%)<br>— Left Lung | 8 | 35.7 (25.0-45.2) | -- | -- | 17.7 (13.0-31.8) | 0.016 |
| **Chest X-ray semi-quantification** |  |  |  |  |  |  |
| Lesion ratio (%)<br>— Right Lung | 12 | 48.5 (30.7-71.8) | 34.0 (23.4-50.7) | 0.024 | 26.7 (4.6-36.4) | 0.005 |
| Lesion ratio (%)<br>— Left Lung | 12 | 47.6 (39.4-72.8) | 39.8 (30.8-49.6) | 0.077 | 28.1 (18.8-55.2) | 0.052 |

\*Chest radiological imaging quantification results are based on available images. Chest CT images on baseline and day 7 post-bevacizumab treatment were from 6 Chinese patients and 2 Italian patients. Chest X-ray images on baseline, day 3 and 7 post-bevacizumab treatment were from 12 Italian patients. Patient No. 15, 19, 21, 22, and 23 were unable to receive chest radiological examination on required timepoints, although they recovered well (shown in Fig. 2). Patient No. 26 was discharged at day 4. IQR = interquartile range.
<sup>†</sup>Wilcoxon matched-pairs signed-rank test.

**Figure 3.**
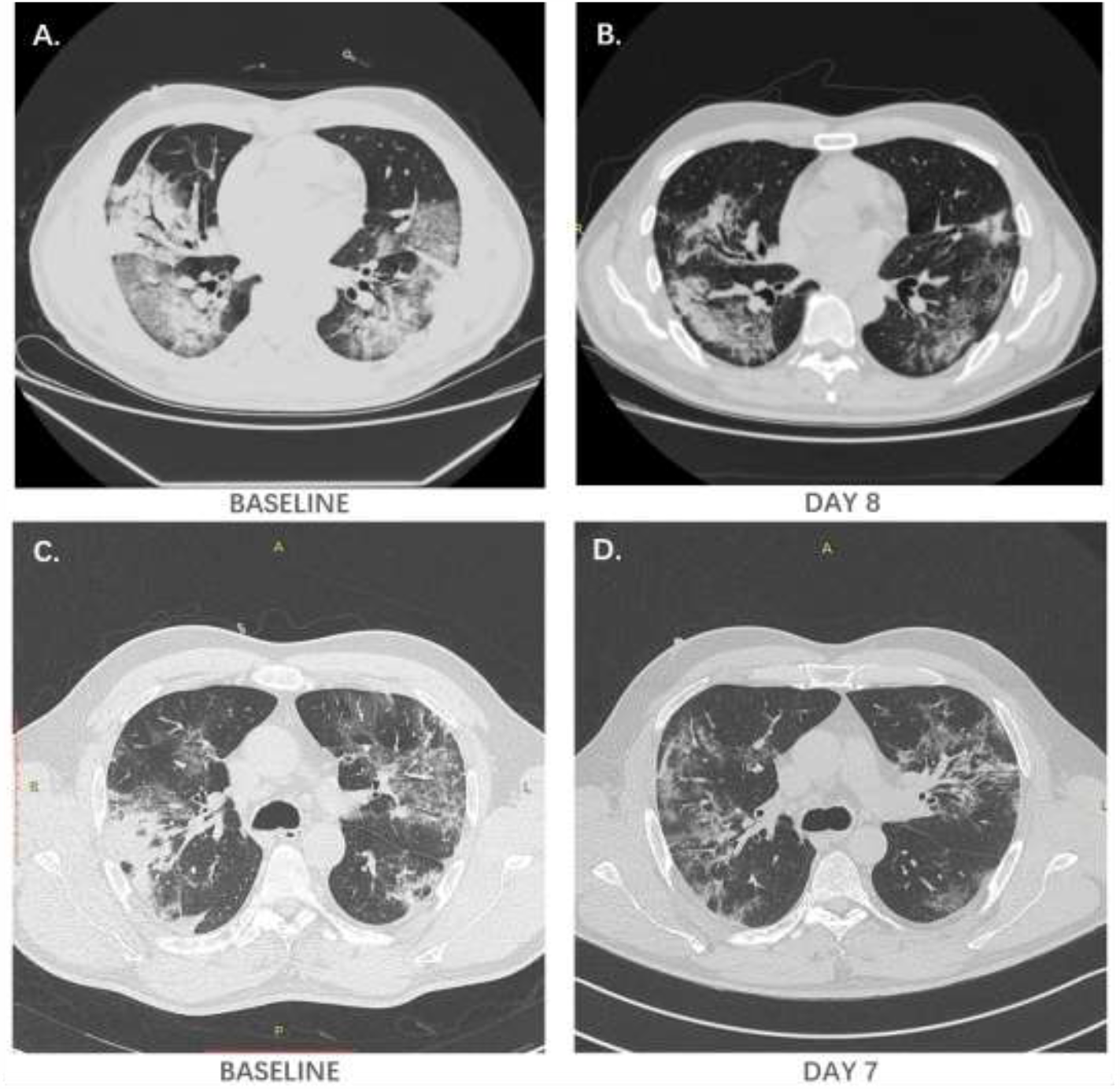
Representative chest CT images. **A.** and **B.** Typical chest CT images of a Chinese patient with severe Covid-19 who received bevacizumab at day 8 relative to the prior treatment baseline. **C.** and **D.** Typical chest CT images of an Italian patient with severe Covid-19 who received bevacizumab at day 7 relative to the prior treatment baseline.

**Figure 4.**
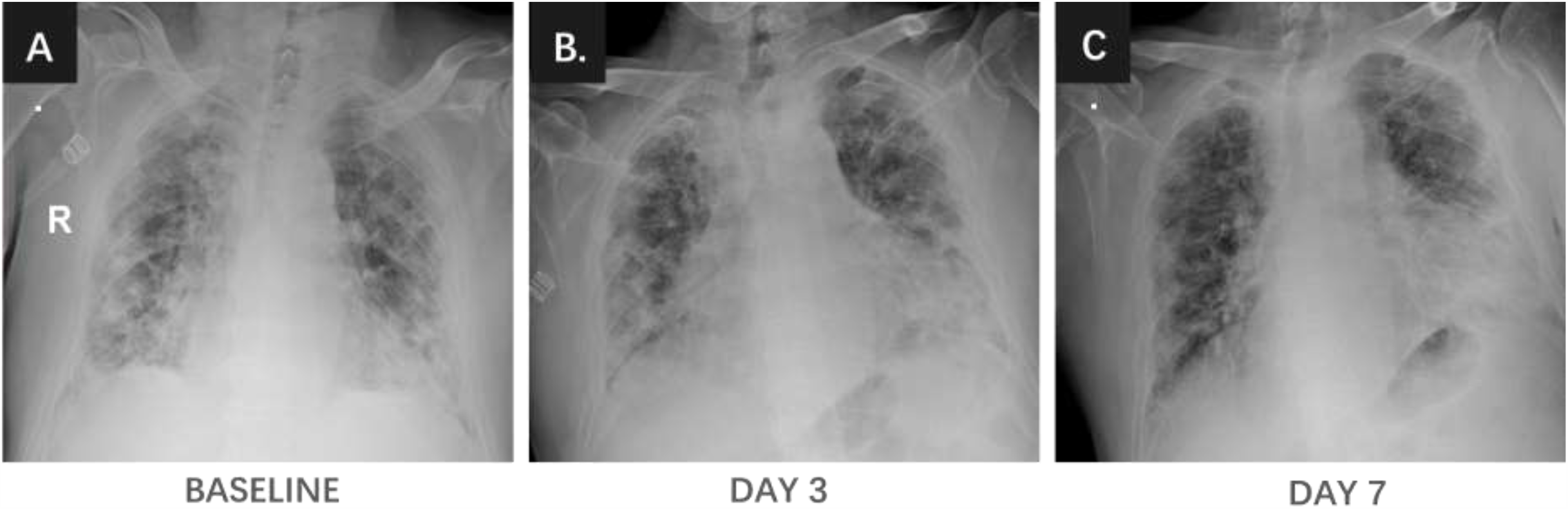
Representative chest X-ray images. **A., B.** and **C**. Chest X-ray images of an Italian patient with severe Covid-19 who received bevacizumab treatment at days 3 and 7 relative the prior treatment baseline.

### Fever Symptom

Surprisingly, we observed a phenomenon of rapid abatement of fever in 13 of 14 febrile patients (93%) within 3 days after bevacizumab treatment regardless of the days of febrile from admission to bevacizumab treatment, which were in a range of 0 to 8 days. One trachea intubation patient was an exception with a persistent fever, who had got sepsis that was evidenced by blood and urine bacteria culture (Fig. 5). From the available values of laboratory tests at baseline and day 7, the peripheral blood lymphocyte counts and the level of CRP were significantly increased and decreased at day 7, respectively (Fig. 5 and Table S1).

**Figure 5.**
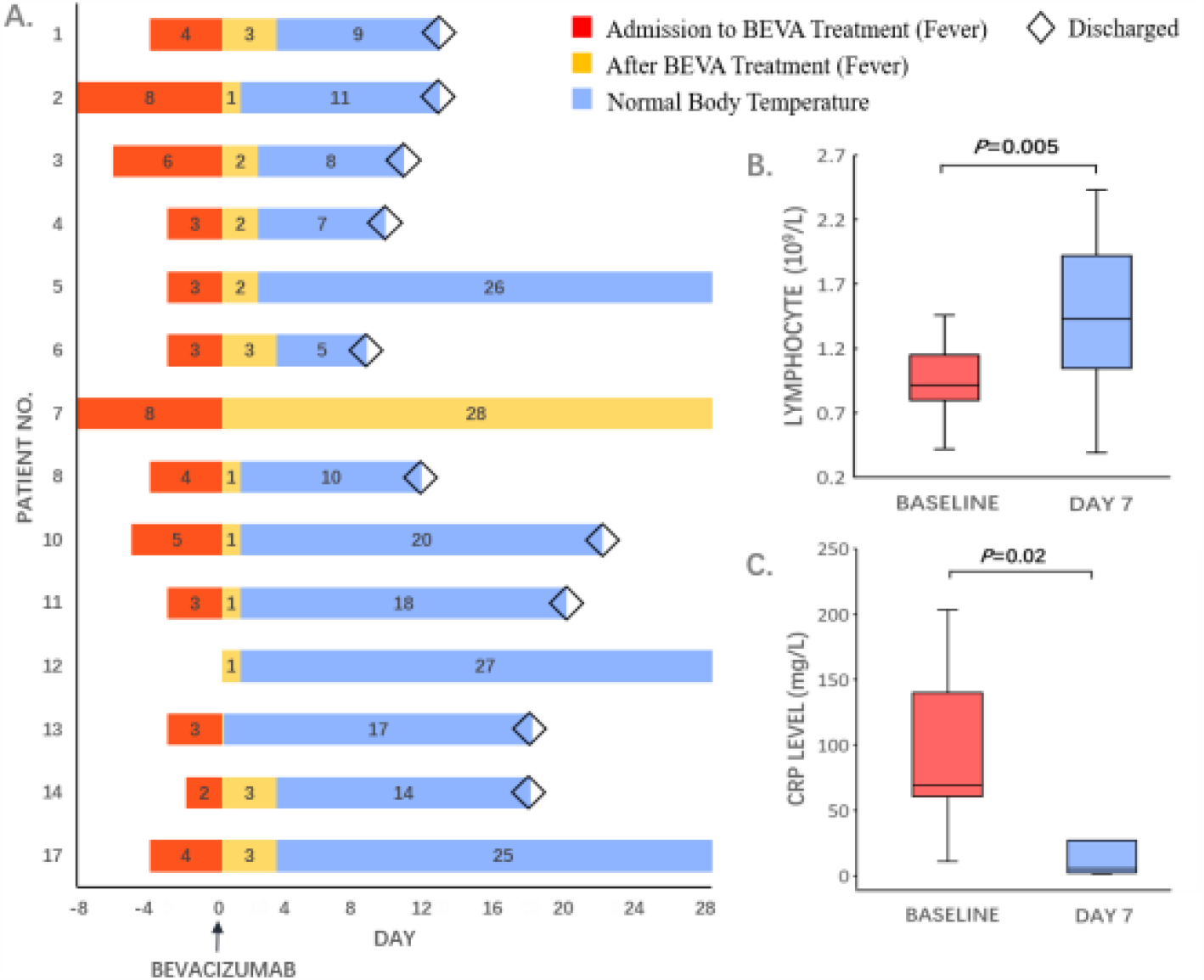
Changes in fever symptom, lymphocyte counts, and inflammation markers of individual patients. **A.** Day 0 marks bevacizumab treatment time point. Final fever statuses are on the discharge date or at the end of 28-day follow-up. Red color columns indicate the fever status prior to bevacizumab treatment. Orange color marks the fever duration after bevacizumab treatment. Blue color represents post-bevacizumab normal body temperature. Diamonds indicate the discharged patients. Patient numbers match the patient numbers in Fig. 2. **B.** Lymphocyte counts at day 7 after bevacizumab treatment relative to the baseline (*p*<0.005). Lymphocyte counts were analyzed from 10 Italian patients and 4 Chinese patients. **C.** C-reactive protein (CRP) analysed at day 7 compared to the baseline (*p*<0.02). CRP values were collected from 8 Italian patients and 1 Chinese patient.

### Safety

Safety data were collected from all 27 patients. Elevation of hepatic enzymes (30%) including alanine aminotransferase and aspartate aminotransferase was the most common adverse event. Other adverse events: 5 patients (19%) showed reduced hemoglobin values; 4 patients (15%) with decreased platelet counts; 3 patients (11%) showed modest elevation of blood pressure, a common adverse event related to bevacizumab; 2 patients (7%) had elevated blood urea nitrogen and normal serum creatinine values; 2 patients (7%) developed sepsis. Haemorrhagic urea, diarrhea, skin rash, muscle pains in lower extremity, superficial phlebitis at the site of venepuncture, and sinus tachycardia with atrial premature beats occurred in one patient, respectively (Table 4).

**Table 4.** Summary of adverse events*

| Events | Chinese<br>(N=13) | Italian<br>(N=14) | Total<br>(N=27) |
| --- | --- | --- | --- |
| Number of patients (%) |  |  |  |
| Adverse events |  |  |  |
| Elevated hepatic enzymes | 4 (31%) | 4 (29%) | 8 (30%) |
| Decreased hemoglobin | 3 (23%) | 2 (14%) | 5 (19%) |
| Reduced platelet counts | 4 (31%) | 0 (0%) | 4 (15%) |
| Elevated blood urea nitrogen | 0 (0%) | 2 (14%) | 2 (7%) |
| Skin rash | 1 (8%) | 0 (0%) | 1 (4%) |
| Local superficial phlebitis | 0 (0%) | 1 (7%) | 1 (4%) |
| Sepsis | 1 (8%) | 1 (7%) | 2 (7%) |
| Oncology bevacizumab-related adverse effects |  |  |  |
| Hypertension (grade ≥2) † | 2 (15%) | 1 (7%) | 3 (11%) |
| Muscle pains in lower extremity | 1 (8%) | 0 (0%) | 1 (4%) |
| Diarrhea | 1 (8%) | 0 (0%) | 1 (4%) |
| Cardiovascular events | 1 (8%) | 0 (0%) | 1 (4%) |
| Hematuria | 1 (8%) | 0 (0%) | 1 (4%) |

\*Adverse events were analyzed for bevacizumab-treated patients whose adverse event information was submitted.
†Hypertension of grade $\geq 2$ referred to recurrent or continuous hypertension for more than 24 hours or symptomatic increase in blood pressure by more than 20 mmHg (diastolic) or to more than 150/100 mmHg if the blood pressure was previously within the normal range.

### External controls

For an external comparison cohort, we screened patients with severe Covid-19 who were eligible as control cases admitted during the same period as the bevacizumab-treated group in two centers, and 26 external control patients were included as a 1:1 ratio to bevacizumab-treated patients. The baseline characteristics were comparable between the two groups (Table 5). Compared with the control group, bevacizumab treatment significantly elevated PaO_2_/FiO_2_ values at days 1 and 7. Standard care had no impact on improving PaO_2_/FiO_2_ values on average in control groups during the comparable period. Relative to controls, the beneficial effect of bevacizumab on oxygenation became obviously significant after 24-hour bevacizumab treatment. These findings indicate that clinical benefits of bevacizumab treatment occur already at early time points (Table 6). Compared with the control group, a significantly increased number of patients improved the 28-day oxygen-support status (control, 62% vs. bevacizumab, 92%) and the discharge rate (control, 46% vs. bevacizumab, 65%) (Fig. S1). The bevacizumab-treated patients also showed less deterioration relative to the control group (control, 19% vs. bevacizumab, 0%). Additionally, bevacizumab treatment significantly shortened the duration of oxygen-support (control, median 20 [IQR 14,28] days vs. bevacizumab, median 9 [IQR 5,19] days, *p*=0.003). When compared our data with other published studies (Table S2), bevacizumab showed potential comparably competitive strength to remdesivir, lopinavir–ritonavir, and convalescent plasma. Thus, our findings are encouraging and may become a first-line therapeutic regimen for treating patients with severe Covid-19.

**Table 5.** Comparison of baseline demographic and clinical characteristics of non-bevacizumab-treated control and bevacizumab-treated group

| **Characteristic** | **Control group**<br>**(N=26)** | **BEVA group**<br>**(N=26)** | ***p* value<sup>†</sup>** |
| --- | --- | --- | --- |
| Age, median (IQR) — yr | 65 (58,72) | 62 (55,66) | 0.131 |
| Maximum body temperature, median (IQR) — °C | 38.4 (37.8,38.8) | 38.5 (38.0,39.0) | 0.223 |
| Symptom onset to admission, median (IQR) — days | 10 (6,15) | 10 (7,15) | 0.887 |
| Sex |  |  | 0.532 |
| Female | 8 (31%) | 6 (23%) |  |
| Male | 18 (69%) | 20 (77%) |  |
| Medical History |  |  |  |
| Heart disease | 0 (0%) | 2 (8%) | 0.490 |
| Hypertension | 9 (35%) | 13 (50%) | 0.262 |
| Diabetes | 1 (4%) | 6 (23%) | 0.099 |
| COPD | 4 (15%) | 3 (12%) | >0.999 |
| Onset symptoms |  |  |  |
| Fever | 23 (88%) | 25 (96%) | 0.610 |
| Dry cough | 15 (58%) | 21 (81%) | 0.071 |
| Expectoration | 7 (27%) | 7 (27%) | 1.000 |
| Fatigue | 10 (38%) | 17 (65%) | 0.052 |
| Shortness of breath | 11 (42%) | 19 (73%) | 0.025 |
| Chest distress | 7 (27%) | 5 (19%) | 0.510 |
| Chill | 1 (4%) | 6 (23%) | 0.099 |
| Headache | 2 (8%) | 6 (23%) | 0.248 |

\*Four data were not recorded; †One datum was not recorded.
†Wilcoxon signed-rank test and Chi-square test.

**Table 6.** Comparison of PaO_2_/FiO_2_ ratios between control and bevacizumab-treated group

| **PaO<sub>2</sub>/FiO<sub>2</sub> ratios** | **Control group**<br>**(N=26)** | **BEVA group**<br>**(N=26)** | ***p* value†** |
| --- | --- | --- | --- |
| **All** |  |  |  |
| Baseline | 197.5 (135.0,295.0) | 196.5 (145.0,273.0) | 0.805 |
| Difference between day 1 and baseline | -23.0 (-71.0,5.0)* | 50.5 (4.0,119.0) | <0.001 |
| Difference between day 7 and baseline | -28.5 (-65.0,62.0) | 111.0 (85.0,165.0)† | <0.001 |
| **Chinese population** |  |  |  |
|  | **(N=12)** | **(N=12)** |  |
| Baseline | 267.5 (156.5,368.5) | 275.0 (231.0,339.5) | 0.686 |
| Difference between day 1 and baseline | -33.5 (-86.0,88.0) | 68.5 (7.5,139.0) | 0.019 |
| Difference between day 7 and baseline | 64.5 (-37.5,134.5) | 93.0 (-9.0,201.0)† | 0.518 |
| **Italian population** |  |  |  |
|  | **(N=14)** | **(N=14)** |  |
| Baseline | 181.5 (135.0,230.0) | 151.5 (140.0,181.0) | 0.597 |
| Difference between day 1 and baseline | -21.5 (-43.5,-2.0)* | 35.5 (4.0,95.0) | 0.001 |
| Difference between day 7 and baseline | -37.5 (-74.0,-24.0) | 120.5 (90.0,165.0) | <0.001 |

\*Two data of day 1 PaO<sub>2</sub>/FiO<sub>2</sub> were absent in Italian control group; †One datum of day 7 PaO<sub>2</sub>/FiO<sub>2</sub> was absent because of discharge in BEVA group as mentioned above.
† Wilcoxon signed-rank test.

## DISCUSSION

Despite initiation of numerous trials since the Covid-19 outbreak, almost no effective therapy is available. In this preliminary trial, we report clinical findings in patients with severe Covid-19 from two medical centers of China and Italy. To the best of our knowledge, the concept of implementing bevacizumab for treating patients with severe Covid-19 is completely novel and has not been explored.

Patients receiving a single-dose treatment with bevacizumab improved the oxygen-support status by 92% during 28-day follow-up without causing death, whereas in the external comparison cohort receiving only standard care, the improvement rate of oxygen-support status was only 62%, and the deterioration rate was 19% including 3 death. Bevacizumab treatment significantly shortened the duration of oxygen-support compared with the control cohort.

There are several unexpected findings from this trial: 1) Rapid improvement of the PaO_2_/FiO_2_ values. Within 24 hours after delivery of bevacizumab, the majority of patients with severe Covid-19 showed rapid improvement of the PaO_2_/FiO_2_ ratio; 2) Rapid abatement of fever. Within 72 hours after bevacizumab, 93% of patients exhibited normal body temperature; Although limited laboratory tests were available, the following two findings are still noteworthy: 3) Increase of PB lymphocyte counts. Patients suffered from lymphopenia, and at day 7 after bevacizumab treatment they showed marked increases of PB lymphocytes; and 4) Anti-inflammation. An over 12-fold decrease of CRP was observed in bevacizumab-treated patients.

Rapid improvement of PaO_2_/FiO_2_ values might reflect the anti-vascular leakiness effect by bevacizumab. A similar rapid response for improving visual acuity by anti-VEGF treatment was also seen in patients with wet age-related macular degeneration (AMD).^25^ VEGF likely significantly contributed to Covid-19-induced inflammation. It is known that VEGF mobilizes inflammatory cells to pathological tissues and anti-VEGF would alleviate fever by antagonizing the virus-triggered inflammation.^26,27^ At this time of writing, the mechanism underlying increases of lymphocytes is unknown. It is speculated that blocking VEGF by bevacizumab might affect extravasation and redistribution of lymphocytes. As lymphopenia is one of the key pathological changes in contributing to Covid-19 fatality,^28,29^ increases of the lymphocytic population would reduce the death rates in severe patients.

Bevacizumab at a dose range of 5-15 mg/kg is routinely used in oncology and about 7.5 mg/kg used in our trial was within the lower range. Bevacizumab-related serious adverse events such as gastrointestinal perforation, haemorrhages, and nephrotic syndrome were absent in both bevacizumab-treated Chinese and Italian populations. No severe safety concerns were detected in this drug use cohort.

It is known that circulating VEGF levels were elevated in patients with severe Covid-19 in Wuhan^5^. Owing to the following reasons, we did not measure plasma VEGF levels: 1) the Covid-19 pandemic-associated limitation of feasibility; 2) plasma VEGF levels may not reflect local VEGF production in pulmonary tissues; and 3) most VEGF isoforms binds to heparin and may be sequestered the tissues where they produce.

The limitation of these results is the small size of the cohort. However, significant improvements of multiple clinical parameters in bevacizumab-treated patients suggest that anti-VEGF might benefit for treating patients with Covid-19. In particular, it is worthy to design future trials by combining bevacizumab with other therapeutic modalities such as anti-viral and anti-inflammatory drugs. Given the limited medical supply and available facility in most medical centers, a single injection of bevacizumab might immediately relieve clinical symptoms and early discharge of patients with severe Covid-19. The therapeutic benefits of bevacizumab monotherapy and combination therapy warrant future validation in patients with severe Covid-19 by randomized and placebo-controlled trials.

## Data Availability

The data related to the results in this article will be available one year after publishment.

## ACKNOWLEDGEMENTS

This study was supported by grants from National Key R& D Program of China (2020YFC0846600) and Shandong Provincial Key R&D Program (2020SFXGFY03). The laboratory of Y.C. is supported through research grants from the European Research Council (ERC) advanced grant ANGIOFAT (project no 250021), the Swedish Research Council, the Swedish Cancer Foundation, the Strategic Research Areas (SFO)–Stem Cell and Regeneration Medicine Foundation, the Karolinska Institutet, the Swedish Children’s Cancer Foundation, the Karolinska Institutet Foundation, the Karolinska Institutet Distinguished Professor Award, the Torsten Soderbergs Foundation, the Maud and Birger Gustavsson Foundation, the Novo Nordisk Foundation–Advance grant, and the Knut and Alice Wallenberg’s Foundation.

## References

1 World Health Organization. Coronavirus disease (Covid-19) outbreak. https://www.who.int.

2 Wu Z, McGoogan JM. Characteristics of and Important Lessons From the Coronavirus Disease 2019 (Covid-19) Outbreak in China: Summary of a Report of 72314 Cases From the Chinese Center for Disease Control and Prevention. JAMA. doi:10.1001/jama.2020.2648 (2020 Feb).

3 Yang, X., Y. Yu, J. Xu, et al. Clinical course and outcomes of critically ill patients with SARS-CoV-2 pneumonia in Wuhan, China: a single-centered, retrospective, observational study. Lancet Respir Med. 8 (5), 475–481 (2020).

4 Bhatraju, P.K., B.J. Ghassemieh, M. Nichols, R. Kim, K.R. Jerome, et al. Covid-19 in Critically Ill Patients in the Seattle Region - Case Series. N Engl J Med. 382 (21), 2012–2022 (2020).

5 Huang, C., Y. Wang, X. Li, et al. Clinical features of patients infected with 2019 novel coronavirus in Wuhan, China. Lancet. 395 (10223), 497–506 (2020).

6 Goyal, P., J.J. Choi, L.C. Pinheiro, et al. Clinical Characteristics of Covid-19 in New York City. N Engl J Med. doi:10.1056/NEJMc2010419 (2020 Apr).

7 Cao, B., Y. Wang, D. Wen, et al. A Trial of Lopinavir-Ritonavir in Adults Hospitalized with Severe Covid-19. N Engl J Med. 382 (19), 1787–1799 (2020).

8 Grein, J., N. Ohmagari, D. Shin, G. Diaz, et al. Compassionate Use of Remdesivir for Patients with Severe Covid-19. N Engl J Med. doi:10.1056/NEJMoa2007016 (2020 Apr).

9 Borba MGS, Val FFA, Sampaio VS, et al. Effect of High vs Low Doses of Chloroquine Diphosphate as Adjunctive Therapy for Patients Hospitalized With Severe Acute Respiratory Syndrome Coronavirus 2 (SARS-CoV-2) Infection: A Randomized Clinical Trial. JAMA Netw Open. 3 (4), e208857 (2020).

10 Torres A, Loeches IM, Sligl W, Lee N. Severe flu management: a point of view. Intensive Care Med. 46 (2), 153–62 (2020).

11 Harris C, Carson G, Baillie JK, Horby P, Nair H. An evidence-based framework for priority clinical research questions for Covid-19. J Glob Health. 10 (1), 011001 (2020).

12 Liu Y, Cox SR, Morita T, Kourembanas S. Hypoxia regulates vascular endothelial growth factor gene expression in endothelial cells. Identification of a 5’ enhancer. Circ Res. 77 (3), 638–43 (1995).

13 Marti HH, Risau W. Systemic hypoxia changes the organ-specific distribution of vascular endothelial growth factor and its receptors. Proc Natl Acad Sci U S A. 95 (26), 15809–14 (1999).

14 Kaner RJ, Ladetto JV, Singh R, Fukuda N, Matthay MA, Crystal RG. Lung overexpression of the vascular endothelial growth factor gene induces pulmonary edema. Am J Respir Cell Mol Biol. 22 (6), 657–64 (2000).

15 Lee, C.G., H. Link, P. Baluk, et al. Vascular endothelial growth factor (VEGF) induces remodeling and enhances TH2-mediated sensitization and inflammation in the lung. Nat Med, 10 (10), 1095–103 (2004).

16 Rodriguez-Morales AJ, Cardona-Ospina JA, Gutiérrez-Ocampo E, et al. Clinical, laboratory and imaging features of Covid-19: A systematic review and meta-analysis. Travel Med Infect Dis. 34, 101623 (2020).

17 Guan WJ, Ni ZY, Hu Y, et al. Clinical Characteristics of Coronavirus Disease 2019 in China. N Engl J Med. 382 (18), 1708–1720 (2020).

18 Liu, Q., R.S. Wang, G.Q. Qu, et al. Gross examination report of a Covid-19 death autopsy. Fa Yi Xue Za Zhi. 36 (1), 21–3 (2020).

19 Tian S, Hu W, Niu L, Liu H, Xu H, Xiao SY. Pulmonary Pathology of Early-Phase 2019 Novel Coronavirus (Covid-19) Pneumonia in Two Patients With Lung Cancer. J Thorac Oncol. 15 (5), 700–4 (2020).

20 Mehta P, McAuley DF, Brown M, Sanchez E, Tattersall RS, Manson JJ. Covid-19: consider cytokine storm syndromes and immunosuppression. Lancet. 395 (10229), 1033–4 (2020).

21 Thickett DR, Armstrong L, Christie SJ, Millar AB. Vascular endothelial growth factor may contribute to increased vascular permeability in acute respiratory distress syndrome. Am J Respir Crit Care Med. 164 (9), 1601–5 (2001).

22 Watanabe M, Boyer JL, Crystal RG. Genetic delivery of bevacizumab to suppress vascular endothelial growth factor-induced high-permeability pulmonary edema. Hum Gene Ther. 20 (6), 598–610 (2009).

23 Varga Z, Flammer AJ, Steiger P, et al. Endothelial cell infection and endotheliitis in COVID-19. Lancet. 2020 May 2;395(10234):1417–1418.

24 Ackermann M, Verleden SE, Kuehnel M, et al. Pulmonary Vascular Endothelialitis, Thrombosis, and Angiogenesis in Covid-19. N Engl J Med. 2020 May 21. doi:10.1056/NEJMoa2015432.

25 Scott IU, VanVeldhuisen PC, Ip MS, et al. Effect of Bevacizumab vs Aflibercept on Visual Acuity Among Patients With Macular Edema Due to Central Retinal Vein Occlusion: The SCORE2 Randomized Clinical Trial. JAMA. 317 (20), 2072–87 (2017).

26 Zhang, Y., Y. Lu, L. Ma, et al. Activation of vascular endothelial growth factor receptor-3 in macrophages restrains TLR4-NF-kappaB signaling and protects against endotoxin shock. Immunity. 40 (4), 501–14 (2014).

27 Janela, B., A.A. Patel, M.C. Lau, et al. A Subset of Type I Conventional Dendritic Cells Controls Cutaneous Bacterial Infections through VEGFalpha-Mediated Recruitment of Neutrophils. Immunity. 50 (4), 1069–1083 e8 (2019).

28 Henry BM. Covid-19, ECMO, and lymphopenia: a word of caution. Lancet Respir Med. 8 (4), e24 (2020).

29 Giamarellos-Bourboulis, E.J., M.G. Netea, N. Rovina, K. Akinosoglou, et al. Complex Immune Dysregulation in Covid-19 Patients with Severe Respiratory Failure. Cell Host Microbe. S1931-3128 (20) 30236-5 (2020).

